# Feasibility of using alternative swabs and storage solutions for paired SARS-CoV-2 detection and microbiome analysis in the hospital environment

**DOI:** 10.1101/2020.05.12.20073577

**Authors:** Jeremiah J. Minich, Farhana Ali, Clarisse Marotz, Pedro Belda-Ferre, Leslie Chiang, Justin P. Shaffer, Carolina S. Carpenter, Daniel McDonald, Jack Gilbert, Sarah M. Allard, Eric E Allen, Rob Knight, Daniel A. Sweeney, Austin D. Swafford

**Affiliations:** Marine Biology Research Division, Scripps Institution of Oceanography, University of California San Diego, La Jolla, CA, USA; Division of Gastroenterology, Department of Pediatrics, University of California San Diego, La Jolla, CA, USA; Department of Pediatrics, School of Medicine, University of California San Diego, La Jolla, CA, USA; Division of Infectious Diseases, Department of Pediatrics, University of California San Diego, La Jolla, CA, USA; Center for Microbiome Innovation, University of California San Diego, La Jolla, CA, USA; Division of Biological Sciences, University of California San Diego, La Jolla, CA, USA; Department of Computer Science and Engineering, University of California San Diego, La Jolla, CA, USA; Department of Bioengineering, University of California San Diego, La Jolla, CA, USA; Division of Pulmonary, Critical Care, and Sleep Medicine, Department of Internal Medicine, University of California, San Diego, La Jolla, CA, USA

**Keywords:** COVID-19, SARS-CoV-2, RT-qPCR, Swab, global health

## Abstract

**Background:** Determining the role of fomites in the transmission of SARS-CoV-2 is essential in the hospital setting and will likely be important outside of medical facilities as governments around the world make plans to ease COVID-19 public health restrictions and attempt to safely reopen economies. Expanding COVID-19 testing to include environmental surfaces would ideally be performed with inexpensive swabs that could be transported safely without concern of being a source of new infections. However, CDC-approved clinical-grade sampling supplies and techniques using a polyester swab are expensive, potentially expose laboratory workers to viable virus and prohibit analysis of the microbiome due to the presence of antibiotics in viral transport media (VTM). To this end, we performed a series of experiments comparing the diagnostic yield using five consumer-grade swabs (including plastic and wood shafts and various head materials including cotton, polyester, and foam) and one clinical grade swab for inhibition to RNA. For three of these swabs, we evaluated performance to detect SARS-CoV-2 in twenty intensive care unit (ICU) hospital rooms of patients with 16 COVID-19+. All swabs were placed in 95% ethanol and further evaluated in terms of RNase activity. SARS-CoV-2 was measured both directly from the swab and from the swab eluent.

**Results:** Compared to samples collected in VTM, 95% ethanol demonstrated significant inhibition properties against RNases. When extracting directly from the swab head as opposed to the eluent, RNA recovery was approximately 2-4x higher from all six swab types tested as compared to the clinical standard of testing the eluent from a CDC-approved polyester swab. The limit of detection (LoD) of SARs-CoV-2 from floor samples collected using the CGp or TMI swabs was similar or better than the CDC standard, further suggesting that swab type does not impact RNA recovery as measured by SARs-CoV-2. The LoD for TMI was between 0-362.5 viral particles while PE and CGp were both between 725-1450 particles. Lastly microbiome analyses (16S rRNA) of paired samples (e.g., environment to host) collected using different swab types in triplicate indicated that microbial communities were not impacted by swab type but instead driven by the patient and sample type (floor or nasal).

**Conclusions:** Compared to using a clinical-grade polyester swab, detection of SARS-CoV-2 from environmental samples collected from ICU rooms of patients with COVID was similar using consumer grade swabs, stored in 95% ethanol. The yield was best from the swab head rather than the eluent and the low level of RNase activity in these samples makes it possible to perform concomitant microbiome analysis.

## Background

Since its appearance in early December of 2019, Severe acute respiratory syndrome coronavirus 2 (SARS-CoV-2), the causative agent of coronavirus disease 2019 (COVID-19), has spread to 197 countries resulting in a total of 539,906 deaths and 11,669,259 confirmed cases as of July 8, 2020[1]. As health officials rush to contain the spread of the disease, federal governments are combating the economic fallout, and there is a pressing need to reopen the economies albeit safely, gradually, and in stages. Large-scale testing and contact tracing remain key for controlling viral spread. In addition,; environmental sampling of microbes can support the epidemiologic investigations of disease outbreaks [2,3] and shows promise for monitoring SARS-CoV-2. However, there are supply and cost limitations with the products currently recommended required by the CDC protocol for sample collection supplies. For example, personal protective equipment, swabs and, viral transport medium (VTM), and personal protective equipment (PPE) are being depleted in developed nations like the United States, and are in even shorter supply in resource limited settings including low- and middle-income countries (3). Broad SARS-CoV-2 surveillance requires microbiologic surface fomite sampling protocols, the efficacy of which hinges on requires inexpensive, readily available swabs and collection reagents to support the large sample sizes at geographic scales necessary to inform public health policy., and the growing need for environmental testing will place additional demands on current swab supplies.

The use of the U.S. Centers for Disease Control (CDC)-recommended viral transport media (VTM) places an additional barrier to efficient and safe deployment of screening and sampling measures. VTM maintains viral viability and therefore the CDC recommends that all samples be handled in a biosafety level-2 (BSL-2) laboratory. VTM also contains antimicrobial agents that limit the type of research studies into likely to interfere with downstream assessment of the microbial context of SARS-CoV-2, such as microbial relationships with that may enable new insights into viral susceptibility and resistance as demonstrated by several recent reports [4–6]. Using inactivating sample collection solutions, such as microbiome assay-compatible alcohols, would increase the number of testing laboratories capable of performing SARS-CoV-2 screening, and ameliorate the risks associated with sample transport and processing. Given these considerations, validation of alternative strategies such as self-administered testing using consumer-grade materials and inactivating storage media is urgently needed.

There are aspects of both the swab and the transport media which must be considered when developing a testing procedure for SARS-CoV-2. From a microbiome perspective, the primary concern with using alternative media and consumer-grade materials is the risk of contaminant RNases and/or PCR inhibitors. The presence of these molecules would increase the false negative rate of SARS-CoV-2 RNA by either degrading the virus, or interfering with reverse transcription and quantitative polymerase chain reactions (RT-qPCR) which are the basis for SARS-CoV-2 testing [7]. In addition, the ability to extract the virus from either the swab or the swab eluent must be elucidated. The fixative property of ethanol could result in nucleic acids adhering to swab heads, reducing the ability to measure SARS-CoV-2 RNA from the swab eluent [6]. To fully address these concerns, large screening efforts comparing the recommended and alternative collection methods are needed. However, given the present scale and urgency of the COVID-19 pandemic outbreak, limiting this comparison to a small number of viable options would greatly expedite providing guidance for alternatives to the supply chain this process while minimizing costs. Here we characterize the suitability of detecting SARS-CoV-2 RNA in experimental conditions as well as COVID-19 patient and built-environment samples using viral-inactivating storage solutions and alternative medical-grade and consumer-grade swabs.

## Materials and Methods

### VTM versus EtOH sample comparison

Nasopharyngeal (NP) swabs were collected from COVID-19 positive individuals (n=39) according to CDC guidelines and were stored in viral transport media (VTM) and transported to the lab on dry ice. For comparison, sterile polyester-head, plastic-shaft (‘PE’, BBL Culture swab REF-220135, Becton, Dickinson and Company) were used to collect nares samples by rotating the dry swab head in the nares for approximately 10 seconds from lab members, COVID-19 patients (n=11), or healthcare workers (n=11) in the Hillcrest ICU, and then immediately placed in 95% ethanol (EtOH), and transported to the lab on dry ice. Eluent nucleic acid extractions were performed on 200 μL of the swab eluent (either VTM or EtOH) using the Omega Mag-Bind® Viral DNA/RNA 96 Kit (catalog# M6246-03), which only uses chemical lysis and does not include a bead beating step. For nucleic acid extraction from the swab head, the MagMAX Microbiome Ultra kit (Cat#A42357, Thermo Fisher Scientific) was used. For the direct comparison of SARS-CoV-2 extraction efficiency, we extracted EtOH eluent and swab separately from the same samples of COVID-19 patients (n=24) with approval of the UC San Diego Institutional Review Board under protocols #150275 and #200613.

### RT-qPCR for VTM and 95% EtOH comparison using polyester-tipped plastic swabs

SARS-CoV-2 detection was performed following a miniaturized version of the CDC protocol. Each RT-qPCR reaction contained 4 μL RNA template, 100 nM forward and reverse primers, 200 nM probe, 3 μl TaqPath (catalog# A15299, Thermo), and RNase-free water to a total reaction volume of 10 μl. All primers and probes were ordered from IDT (catalog# 10006606). RT-qPCR was performed on the Bio-Rad CFX384 Touch Real-Time PCR Detection System following the CDC thermocycling guidelines. Serial dilutions of the Hs_RPP30 Positive Control plasmid (catalog# 10006626, IDT) or 2019-nCoV_N_Positive Control plasmid (catalog# 10006625, IDT) were included to extrapolate human RNase P (Rp) and SARS-CoV-2 copy numbers, respectively. The SARS-CoV-2 N1 marker gene was used for detection and quantitation [8] [9].

### Validation of use of alternative swabs (testing inhibition of SARS-CoV-2 detection)

The standard swab type approved for use in SARS-CoV-2 detection is a synthetic fiber swab with a plastic or wire shaft polyester head. In addition to a CDC approved device, we tested an additional five alternative swabs that included both plastic and wood materials for the shaft and polyester, foam, or cotton materials for the swab head. The exact devices used were sterile rayon polyester-head, plastic-shaft (‘PE’, BBL Culture swab REF-220135, Becton, Dickinson and Company); sterile foam-head, plastic-shaft (‘BDF’, Flock PurFlock REF-25-3606-U-BT, Becton, Dickinson and Company); non-sterile cotton-head, plastic-shaft in use by The Microsetta Initiative (‘TMI’, SKU#839-PPCS, Puritan Medical Products); non-sterile cotton-head plastic-shaft consumer-grade (‘CGp’ Part #165902, CVS Caremark Corp.); non-sterile cotton-head wooden-shaft consumer-grade (‘CGw’, Part#858948, CVS Caremark Corp.); and non-sterile cotton-head, wooden-shaft (‘Pu’, REF-806-WC, Puritan Medical Products). The goal was to evaluate if detection of SARS-CoV-2 was reduced with certain swab types from both the eluent (standard protocol) and swab head directly (new method). A total of six swab types were compared and All swabs were processed following the standard SARS-CoV-2 protocol provided by the CDC [6]. The six swab types were used: sterile rayon polyester-head, plastic-shaft (‘PE’, BBL Culture swab REF-220135, Becton, Dickinson and Company); sterile foam-head, plastic-shaft (‘BDF’, Flock PurFlock REF-25-3606-U-BT, Becton, Dickinson and Company); non-sterile cotton-head, plastic-shaft in use by The Microsetta Initiative (‘TMI’, SKU#839-PPCS, Puritan Medical Products); non-sterile cotton-head plastic-shaft consumer-grade (‘CGp’ Part #165902, CVS Caremark Corp.); non-sterile cotton-head wooden-shaft consumer-grade (‘CGw’, Part#858948, CVS Caremark Corp.); and non-sterile cotton-head, wooden-shaft (‘Pu’, REF-806-WC, Puritan Medical Products). To evaluate if the raw swab materials had any background contaminants such as RNase, which would decrease the sensitivity, we added 600 ng of purified, DNA-free human lung RNA (Cat#AM7968, Thermo Fisher Scientific) onto each of the six swab types in triplicate and immediately stored in two storage solutions (500 μL 95% EtOH and 500 μL 91% isopropanol). Separately, two sets of six, 10-fold serial dilutions of human RNA were included as controls directly.

The same quantity of human RNA (600 ng) along with an equal volume (5 μL) of SARS-CoV-2 RNA was added to either 95% EtOH (n=3) or 91% isopropanol (n=3) in the presence of 0, 2.5, and 25 μg RNaseA (in triplicate) to assess any inhibition offered against RNase contaminants. Four negative (swab only) and four positive (swab + 600 ng spiked human RNA + 5 μL spiked SARS-CoV-2 RNA [~20,000 copies per μL]) controls were included.

### Limit of detection comparison of swabs using floor as substrate

To estimate the limit of detection and compare the viral yield across three swab types (PE, CGp, and TMI), a serial dilution of viral particles was spiked onto floor swabs. In brief, separate 25 cm x 25 cm areas of the floor from a low-traffic common room inside a building with no SARS-CoV-2 research activities (i.e., Marine Biology research building at UC San Diego) were swabbed with a total of 24 swabs per swab type. Swabs were processed in groups of six by swabbing a quarter of that 625-cm^2^ space, with each swab ultimately covering an *ca*. 26-cm^2^ area, the similar surface area (25 cm^2^) used for detection of low biomass samples in JPL spacecraft assembly clean rooms based on previous work in the JPL spacecraft assembly facility [10]. Swabs were then stored at room temperature for *ca*. 1 hr in a 2-mL deep-well 96-well plate during transport back to a BSL-2 laboratory at UC San Diego. A single serial dilution of SARS-CoV-2 viral particles [BEI Resources: Cat# 52286, Lot# 70033548] was made at the following concentrations: 232000, 2320, 1160, 580, 290, 145, and 72.5 viral particles per μL. A total of 5 μL of each dilution, or water as a negative control, was pipetted onto each swab type in triplicate and then immediately placed into 95% EtOH. Swabs in EtOH were then stored overnight at -80°C until processing. Upon processing, an additional 24 ‘no swab’ controls were included whereby 5 μL of the dilutions were dispensed directly into the extraction plate lysis buffer. Samples were processed using the same nucleic extraction method as described for swab heads above, and eluted in 75 μL of elution buffer. For RT-qPCR, 5 μL of template was used for each marker N1 and Rp. To address potential issues of non-normality, total copies were compared across swab types at each individual dilution using Kruskal-Wallis tests with Benjamini-Hochberg FDR 0.05 post-hoc test.

### Patient and hospital environmental sampling

All study patients were hospitalized with clinical concerns for COVID-19 and received standard diagnostic testing. Study samples were collected from subjects’ nares or hospital surfaces using three dry swab types (PE, TMI, CGp) under the UC San Diego Institutional Review Board protocol #150275 and #200613. Both nasal samples and hospital surfaces were collected using three dry swab types (PE, TMI, CGp). Nasal samples were collected by inserting the swab into one nostril to the depth of approximately 2-3 cm and rotated for 5–10 seconds. Hospital surfaces sampled included the floor inside the patient’s room (*ca*. 625-cm^2^ area) and the patient’s bedrail. All swabs were immediately placed in a collection tube containing 0.5-1.0 mL 95% EtOH, stored on dry ice, and processed for RNA or total nucleic acid extraction (Supplementary Methods).

### Extraction and RT-qPCR of hospital swabs and controls

All swab comparison-and hospital samples were processed according to the manufacturer’s protocol using the MagMAX Microbiome Ultra kit (Cat#A42357, Thermo Fisher Scientific), and eluted into 70 μL buffer. For RT-qPCR, 5 μL sample was processed using the standard SARS-CoV-2 protocol provided by the CDC (Cat# 2019-nCoVEUA-01[11]).

### Microbiome processing and analysis

A subset of 40 samples were processed for 16S rRNA sequencing using established EMP protocols (https://earthmicrobiome.org/protocols-and-standards/16s/). These included 18 floor samples, 21 nasal samples, and 1 negative control. Floor samples included all triplicates from the three swab types (PE, TMI, and CGp) from two patient rooms (patient 7 and 18). The nasal samples included triplicates of all three swab types from patient 1, triplicates of PE and CGp from patient 7, and triplicates of PE and TMI from patient 18. The same previously extracted nucleic acid template, which was concurrently used for RT-qPCR, was used as template for 16S rDNA library generation (amplifying the DNA). Specifically, 0.4 μL of nucleic acid was processed in 10 μL 16S rRNA PCR reactions following the miniaturized protocol [12] using the 515f/806r EMP primers, and sequenced on an Illumina MiSeq [13-16]. Samples were then processed in Qiita (Study ID 13275) [17] and analyzed using the QIIME2 [18,19] [version?] pipeline with Deblur [20] 1.1.0 as the method of sOTU generation. Samples were visualized in PCoA plots in Qiita using EMPeror [21]. Beta diversity was calculated using Unweighted Unifrac and compared with PERMANOVA (999 permutations).

### Statistics and visualizations

Visualizations and statistical comparisons performed using PRISM 8.0 and the limit of detection determination were consistent with CDC recommendations whereby samples with a Ct value greater than 40 were omitted [8].

## Results

Our experimental design sought to answer three primary questions: whether the efficacy of SARS-CoV-2 detection is influenced by the following three variables: 1) does the swab storage solution (95% EtOH vs 91% isopropanol) impact the sensitivity of detection; 2) which sample fraction, swab head or eluent, provides better detection fidelity; and 3) does the swab head material type matter? To do this we designed a series of experiments to compare RNA recovery as measured by RT-qPCR using multiple swab types and storage solutions. We additionally did environmental sampling in a hospital environment with a subset of swab types for comparison.

### Feasibility of 95% EtOH for sample storage and extraction from use of swab head rather than eluent

To evaluate the feasibility of switching from VTM to a more readily-available, viral-inactivating sample collection solution, we compared the extraction efficiency of polyester-tipped plastic-shafted nasopharyngeal (NP) swab samples stored in VTM versus nasal samples collected using CDC-recommended polyester-tipped plastic-shafted (PE) swabs stored in 95% ethanol (EtOH). When mirroring the CDC protocol, which calls for extraction from 200 μL of the eluent from VTM surrounding NP swabs, we had significantly lower recovery of human RNA in 95% EtOH eluent compared to VTM (Figure 1a; one-way ANOVA with Tukey’s multiple comparison, VTM eluent vs. EtOH eluent p<0.001). However, similar levels of human RNA were recovered when extracting from the EtOH-preserved swab head itself (Figure 1a; one-way ANOVA with Tukey’s multiple comparison, VTM vs. EtOH swab p=0.3). In a subset of seven COVID-19 patient nares samples stored in 95% EtOH, we also detected significantly higher SARS-CoV-2 viral load in RNA extracted from the swab head versus eluent (Figure 1b; one-tailed paired Student’s *t*-test p=0.03).

**Figure 1.**
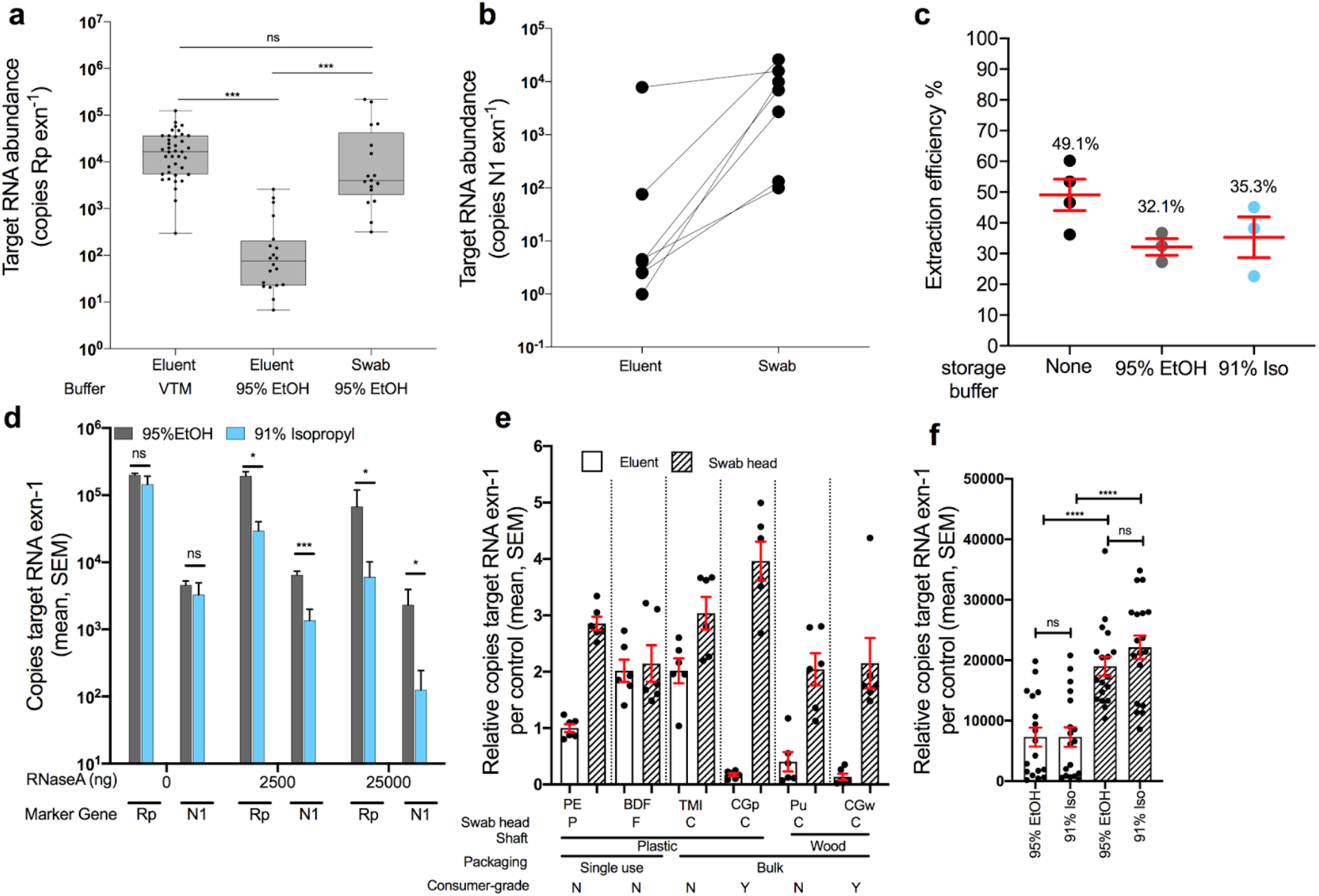
Validation of alternative swabs and storage buffer (95% EtOH and 91% isopropanol) in RNA recovery and detection of COVID-19. **a)** Human RNAse P gene (Rp) amplification was used to compare nucleic acid extraction efficiency across sample processing methods. Clinical gold-standard polyester-tipped plastic-shaft NP swabs stored in VTM and extracted from 200 μL of eluent (left, n=39) have significantly higher copy numbers compared to 200 μL EtOH eluent from PE nares swabs (middle, n=22), but not when extracted from the EtOH-preserved swab head (right, n=18). One-way ANOVA with Tukey’s multiple comparison VTM eluent vs EtOH eluent p=<0.001, EtOH eluent vs EtOH swab p<0.001, VTM vs EtOH swab p = 0.266. **b)** Extrapolated viral RNA copy number from COVID-19 positive nares samples collected with BD polyester swabs in the hospital stored in 95% EtOH and extracted from either the eluent or swab from the same sample (n=24, one-tailed paired Student’s T-test p=0.032). **c)** Proportion of RNA recovered across three storage buffers: None, 95% EtOH, and 91% isopropanol using commercial human RNA added to storage buffers (ns, one-way ANOVAp>0.05). **d)** Evaluation of RNaseA inhibition by 95% EtOH (grey) and 91% isopropanol (blue) using either the human Rp or SARS-CoV-2 N1 primer set on control RNA added to each solution (unpaired t-tests of 95% EtOH vs 91% Iso per each marker at 0, 2500, 25000 ng RNaseA). **e)** Comparison human RNA recovery across six swab types (PE=polyester ‘commercial’, BDF=BD foam ‘commercial’, TMI=BD TMI ‘commercial’, CGp=plastic ‘consumer-grade’, Pu=Puritan ‘commercial’, CGw=wood ‘consumer-grade’), extracted from 200μL eluent (blank bar) or the swab head. Recovery for each swab type is normalized to the CDC recommended method (eluent from PE swab). A ‘2’ would indicate there was 2x more RNA recovered whereas a 0.5 would indicate a 50% reduction in RNA recovery. **f)** Total RNA copies per extraction for all samples which are grouped by sample-type (eluent or swab head) and storage buffer (95% EtOH or 91% isopropanol). Pairwise comparisons performed within sample-type (not significant) and across sample-type controlling for storage buffer (Mann-Whitney, U=test statistic).

To more quantitatively determine the effects of alcohol-based preservation media, we extracted RNA from a pure, commercial sample of human RNA added to water, EtOH, or 91% isopropanol, and found no impact on extraction efficiency (Figure 1c; one-way ANOVA, p>0.05). Next, we examined whether alcohol storage solutions had any protective properties of RNA, specifically a possible inhibitory effect on RNases that might be present in the environment. If alcohol inhibits the RNaseA, one would expect to see similar amounts of RNA as without RNaseA added in control experiments. In the presence of abundant RNaseA added to the solution, 95% EtOH protected both human RNA and SARS-CoV-2 RNA better than 91% isopropanol. Only a moderate decrease in total RNA recovery was observed, at the most extreme concentration of 25 mg per reaction, which is equivalent to the standard amount used for RNA removal during DNA extraction (Figure 1d).

### Comparison of alternative swab types against standard CDC approved polyester swab

Given that the performance of eluent vs. swab-based extractions in each alcohol may depend on the swab tip and body composition, we next tested RNA recovery from both the swab head and the surrounding eluent from a range of medical- and consumer-grade swabs (Methods) (Figure 1e). The RNA yield was highest from swab heads compared to eluent regardless of the swab type and whether stored in 95% EtOH (p<0.0001, U=37, Mann-Whitney) or 91% isopropanol (P<0.0001, U=28, Mann-Whitney) (Figure 1e-f). The storage solution did not impact RNA quality (Supplemental Figure 1b, Mann-Whitney, p>0.05), although swab type had a minor impact (Supplemental Figure 1c, Kruskal-Wallis p=0.03, KW=12.17)[22]. To compare impacts of various alternative swabs, we normalized the recovery of each test to PE eluent, indicated by ‘1’ (Figure 1e), which is the standard CDC approved method. Thus, any sample with a value greater than 1 would indicate an enhanced recovery of RNA, whereby less than 1 indicates a lower recovery of RNA compared to the standard. The RNA recovery ratio of swab-to-eluent and total yield varied among swab type (p<0.0001, KW=28.37, Kruskal-Wallis for eluent, and p<0.0001, KW=15.43, Kruskal-Wallis for swab heads) (Supplemental Figure 2). This difference in performance may relate to the differences in observed adsorption capacity across swab types (Shapiro-Wilkes p=0.1, w=0.8357; ANOVA p=0.0001, F=7.5, R^2^=0.56). TMI adsorbed the least (84.5 μL, 20.4; mean, SD) followed by plastic shafts (PE: 141 μL, 23.1; CGp: 143.3 μL, 29.9) (Supplemental Figure 3). CGp swabs had the highest recovery of RNA from the swab head, while TMI swabs had the highest overall recovery of RNA when combining both eluent and direct swab extractions (Figure 1e, Supplemental Figure 2).

**Figure 2.**
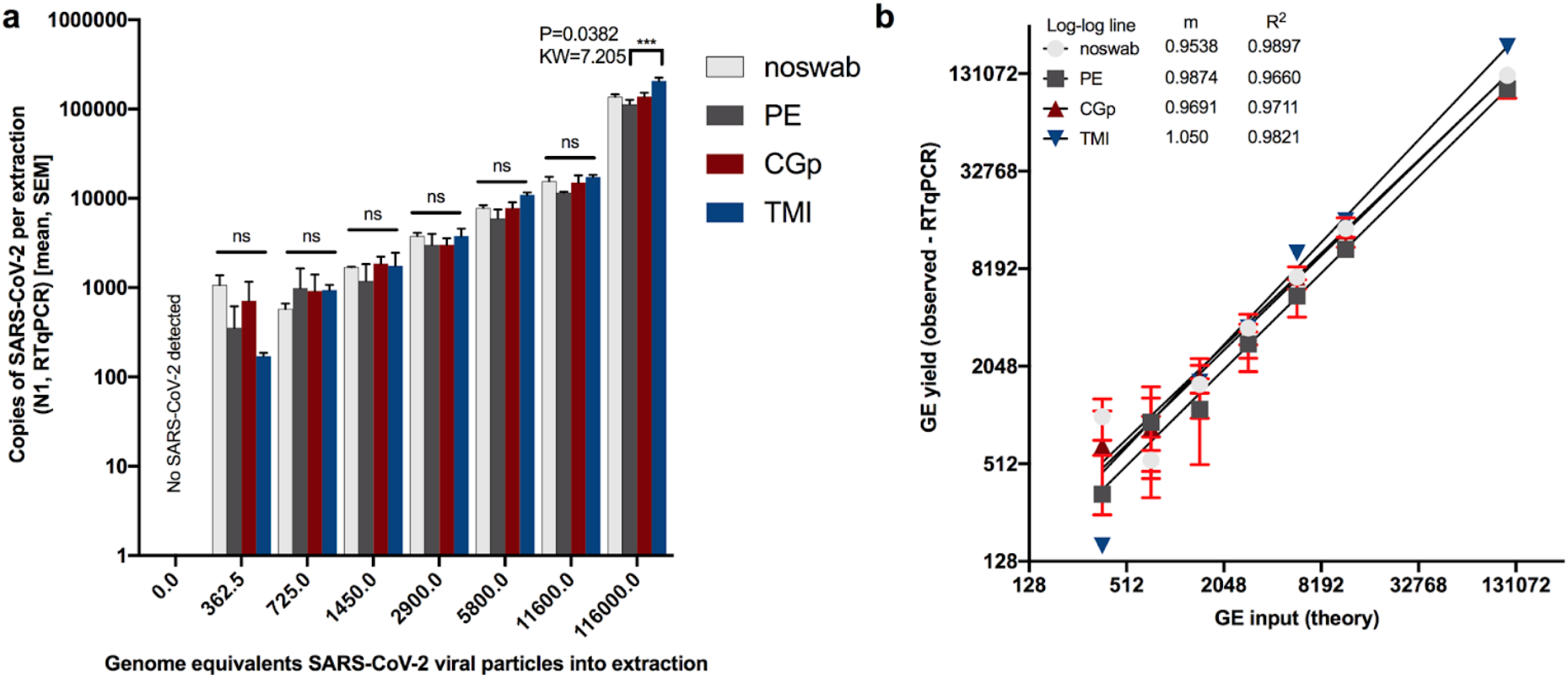
Limit of detection of SARs-CoV-2 viral particles across swab types. (Polyester ‘PE’, CGp, and TMI). “Noswab” refers to direct extraction of viral particles. a) Comparisons of total RNA recovery per extraction across swab types including ‘noswab’ performed at each dilution (Kruskal Wallis test). Comparison of (theory input Genome Equivalents ‘GE’) to measured GE of triplicates (mean, SEM) by RTqPCR of SARs-CoV-2. Non-linear regression analysis of each dilution series for noswab, PE, CGp, and TMI swabs.

### SARSs-CoV-2 limit of detection comparison across swab types

We next assessed whether the swab type used would impact the recovery of SARS-CoV-2 and alter the limit of detection when using non-CDC-recommended swabs (CGp or TMI compared to PE). All negative controls for floor swabs were indeed negative for SARS-CoV-2 using N1 and N2 (Supplemental Table 1, Figure 2) and all ‘no-swab’ controls which only had SARS-CoV-2, were negative for human Rp (Supplemental Table 1). For the ‘no-swab’ and TMI swab, SARS-CoV-2 was detected in all of the three replicates at the lowest input of 362.5 genome equivalents ‘GE’, whereas the lowest dilution for all three replicates to be positive for CGp and PE swabs was 1450 GE (Supplemental Table 1, Figure 2a). This suggests the limit of detection for neat and TMI swabs is likely between 0 and 362.5 GE per reaction whereas both CGp and PE were less sensitive with an expected limit between 750 and 1450 GE per reaction. There was a strong correlation between the input or theoretical GE and the measured GE with slopes all greater than 0.95 and the R^2^>0.96. Despite TMI appearing to have the best overall performance in SARSs-CoV-2 detection followed by PE and then CGp, the total viral yield did not differ across swab types at the lowest dilution of 362.5 (P>0.05, Kruskal-Wallis test) (Figure 2a). Specifically, multiple post-hoc comparisons showed that variation across swab-type only existed at the highest concentration (116,000 GE) with the TMI swabs having a higher viral recovery compared to PE swabs (P=0.04, KW=7.21) (Figure 2). Rp yield was also compared across swab types and across viral inputs to characterize the variation in input biomass. For each swab type, human Rp gene was equally detected across the titrations indicating the swab method was sufficiently controlled (Supplemental Figure 4a). Swab type, however, did suggest that the Rp gene was highest in the PE swab as compared to the CGp and TMI swabs (Kruskal-Wallis: P<0.0001, KW=41.41) (Supplemental Figure 4b). This result suggests that PE swabs may adsorb more biomass. However, when we compared the variation in Cq values of hospital samples of nares and floor from the same hospital using PE swabs, we observed Rp values that varied over six orders of magnitude (Supplemental Figure S5), much greater than the three orders of magnitude observed across swab types. Specifically, for floor samples, the Rp yield (copies per extraction) range across swab types was 149-3368 copies for PE, 0-3980 for CGp, and 0-207 for TMI.

### Hospital proof of concept study

Based on the results from these initial experiments, we conducted a proof-of-concept study in the clinical setting by performing RT-qPCR for the SARS-CoV-2 N1 amplicon and human RNase P gene on RNA extracted from the swab head of nasal samples collected using TMI and/or CGp swabs alongside the recommended PE swabs. Of the 20 participants sampled, 16 tested positive for SARS-CoV-2 at admission and were designated as COVID-19(+). The average time from diagnosis to sampling was *ca*. 4.2 days, with a NP swab test occurring within 72 hours of the time of nasal sampling. Of the 12 nasal samples using the PE swab preserved in EtOH from COVID-19(+) patients, nine were positive for the presence of SARS-CoV-2 or a false negative rate of 25% (Figure 3a) compared to 14/16 SARS-CoV-2 positive NP swabs for the same group of patients, a false negative rate of 12.5%. For CGp and TMI swabs, 8/12 and 5/10 were positive for nares, respectively (Figure 3a). These rates of false negatives are similar, as compared to the 37.5% false negative rate reported for plastic-shafted polyester-tipped nasal swabs collected in VTM and extracted from the eluent (Wang et al. 2020). As the degree of viral shedding is known to vary over the course of the disease [23], we compared the performance in the subset of COVID-19(+) patients with an NP-positive swab result within 72 hours of the time of sampling, and observed reduced false negative rates of 18.2% (PE), 25% (TMI), and 30% (CGp). We next compared success rates across swab samples from the built environment. On the floor samples, the CGp had the highest success rate at 75% in detection of SARSs-CoV-2 from SARSs-CoV-2 positive patient rooms whereas PE detected SARSs-CoV-2 in 63% of rooms, and TMI in 44% of rooms (Figure 3a). Bedrail samples had the lowest frequency of detection, 5/16 (31%), for each swab type (Figure 3a). For SARSs-CoV-2 negative patients admitted to the same hospital for other reasons, all nares and bedrail samples were negative, whereas one floor sample using the PE swab detected SARSs-CoV-2 (Figure 3b).

**Figure 3:**
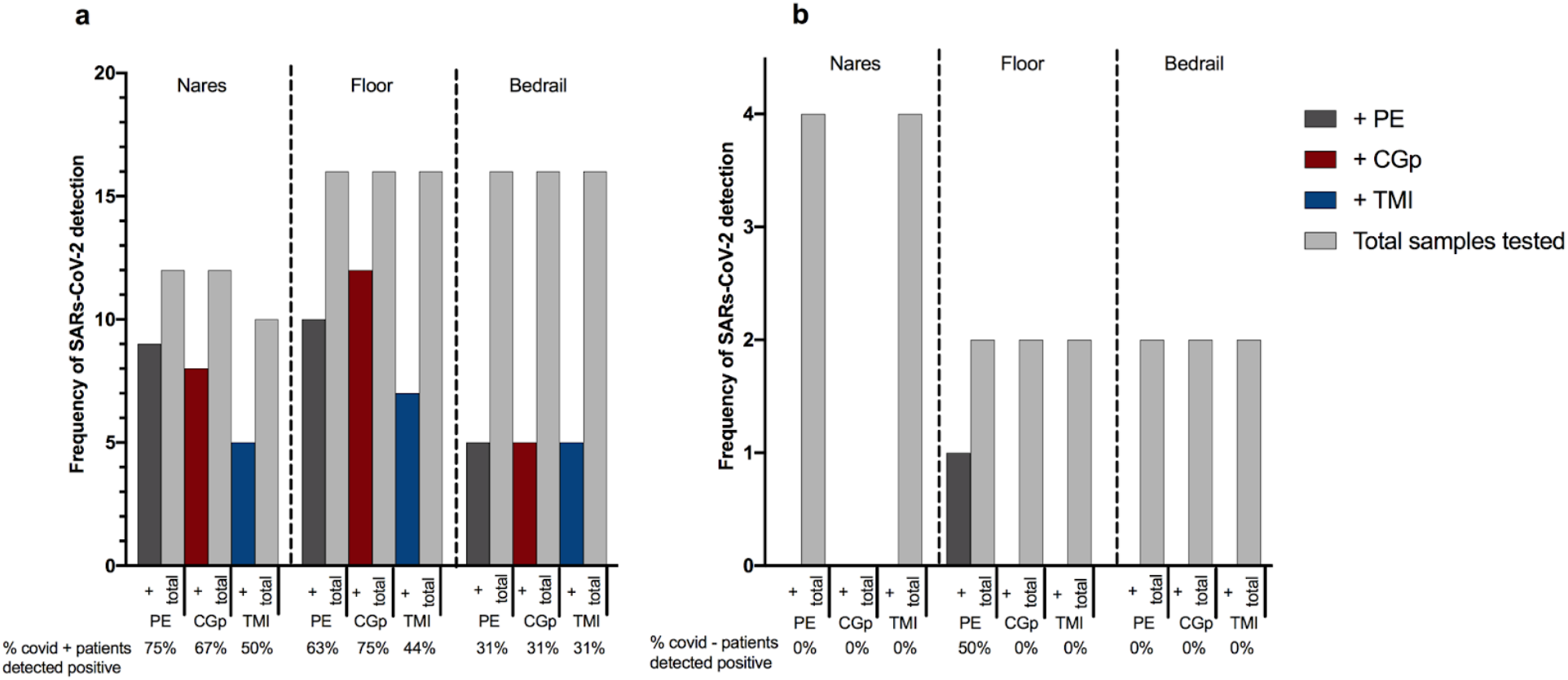
Comparison of CDC approved PE swabs, consumer-grade CGp and bulk TMI swab congruence compared to clinical-grade hospital tests using polyester-tipped plastic shafted NP swabs for twenty participants in the clinical setting. a) SARs-CoV-2 positive patients (n=16) sampled with three swab types across three environments: nares, floor, and bedrail. ‘+’ samples (dark grey = PE, red = CGp, blue = TMI) refer to samples which tested positive for SARs-CoV-2 out of the total samples tested for that particular swab type (light grey bar). Percentage of positive tests per swab type are below x axis for each environmental sample. b) SARs-CoV-2 negative patients (n=4) with three swab types across three environments: nares, floor, and bedrail. Same nomenclature as above.

The observed differences in detection among nares and environmental samples, taken in context of results that Since our previous experiment demonstrates that suggest swab type does not impact SARSs-CoV-2 detection, this suggests that variation in sample collection from the nares and other environmental samples has an important role in detection sensitivity. When swabbing an environmental surface or body site (i.e., nares), there is inherent variation in the swabbing event which can be attributed both to stochastic differences in biomass (human cells, dust, etc.) and the overall assay (nucleic acid extraction and RT-qPCR). To evaluate if certain sampling locations or swab types were more variable than others, we calculated the intra-assay coefficient of variance (CV) of the Cq values. The CV was significantly higher in patient nasal samples compared to control (RNA spike-in) samples (P=0.0018), with a median difference in variance of 2.5 (Supplemental S6a). Swab types also demonstrated an effect with CGp and PE differences being significant as compared to the control (P=0.0012) (Supplemental Figures S6b).

### Microbiome analysis

To determine the feasibility of co-opting nucleic acid for microbiome processing, we processed a subset of samples (n=40) spanning a total of three patients, two sample types (floor and nasal) and the three swab types. After processing with Deblur, the total number of reads per sample were compared (Figure 4a). Read counts were highly variable across sample types and for each patient but were consistent within the swab types for each comparison. For floor samples in patient room 18, PE swabs had the highest number of reads followed by TMI and CGp. For nasal samples however, patient 1 had the higher read counts from TMI while patient 7 and 18 both showed slightly lower read counts for PE swabs as compared to alternative swabs. The differences were minor however and are primarily differentiated by patient room (Figure 4a). After rarifying to 5000 reads a PCoA plot was generated from using Unweighted UniFrac distances (Figure 4b). Samples which were collected using different swab types clustered together when controlling for patient room and sample type, suggesting indicating that the swab type used does not have a negative impact on microbiome analysis (Figure 4b). When analyzing all samples together, sample_type (floor vs nasal) and patient number (7 vs 18) were both significant drivers of the microbiome community (sample_type: PERMANOVA n=24, group=2, P=0.001, Fstat=6.94; patient_num PERMANOVA n=24, group=2, P=0.001, Fstat=6.92) whereas swab type did not have an effect (P=0.164). Distances between swab types were lower than distances between patients for both floor (Supplemental Figure S7a) and nasal (Supplemental Figure S7b) samples, with patient 7 exhibiting higher variation than patient 18. Floor samples generally had a higher microbial diversity compared to nasal swabs, with *Staphylococcus*, *Corynebacterium, Pseudomonas, Streptococcus*, and Enterobacteriaceae being the more dominant taxa. Nasal samples however were mostly enriched by either *Staphylococcus* or *Corynebacterium*, with patient 7 having a higher abundance of *Lawsonella* (Figure 4c).

**Figure 4.**
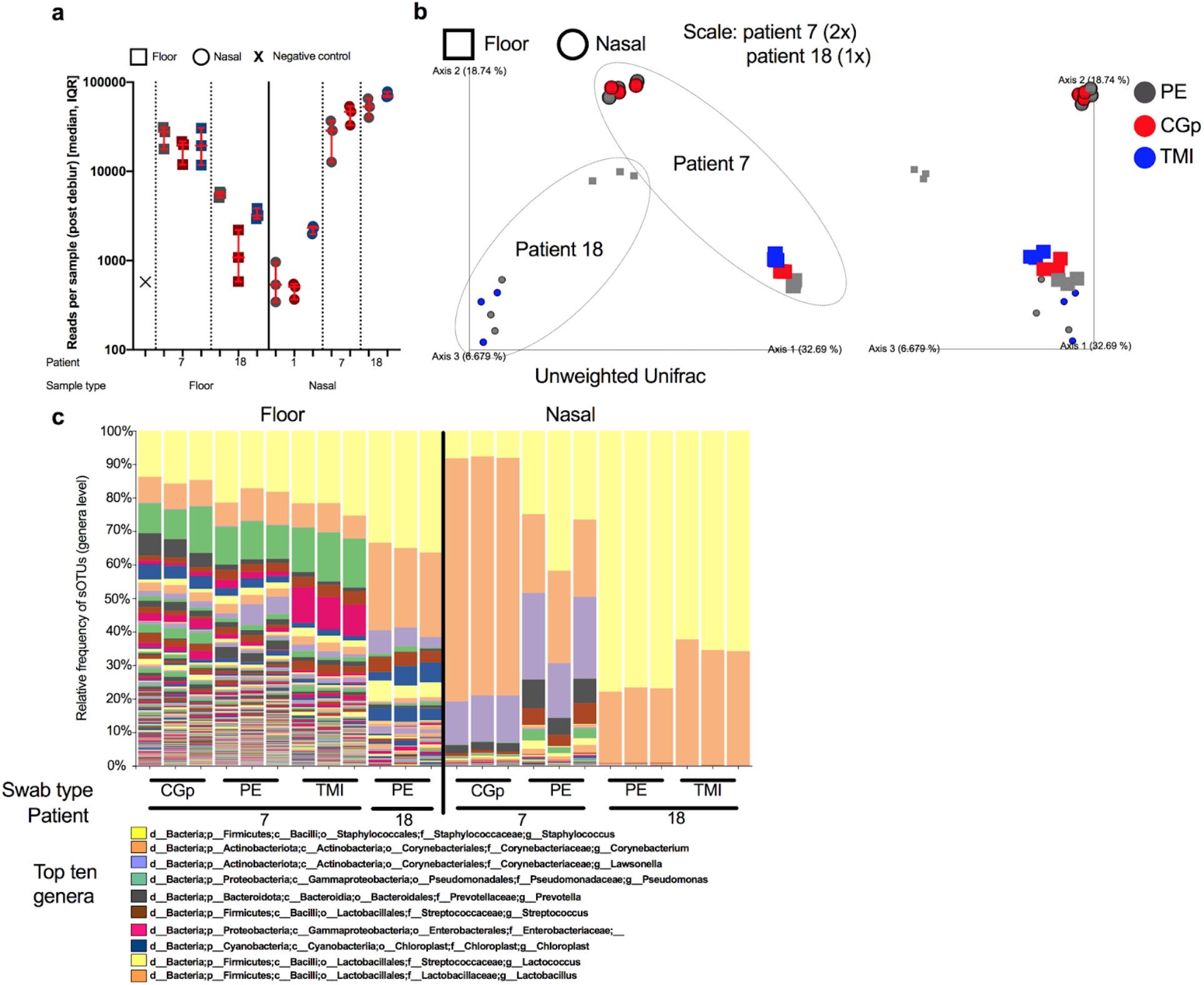
Microbiome 16S rRNA sequencing validation across sample types, patients, and swab types. a) Total number of reads per sample (40 samples sequenced) after processing through deblur pipeline stratified by sample type (floor ‘square’ vs nasal ‘circle’), patient number (1, 7, and 18) and colored by swab type (PE = grey, CGp = red, TMI = blue). Error bars represent median, IQR for triplicate biological replicates per sample. b) Unweighted UniFrac PCoA plot of samples rarified to 5000 reads. Enlarged samples (2x) indicate patient 7 whereas (1x) indicates patient 18. Swab types are colored (PE = grey, CGp = red, TMI = blue) and shapes (floor ‘square’ vs nasal ‘circle’) indicate sample type. Grey dotted line goes around each patient. c) Stacked bar plot collapsed at genera level with top ten abundant genera labeled in the legend.

## Discussion

When assessing whether it will be possible to adapts and switch collection methodology to enable more affordable, more widely available, and more inter-assay compatible collection methods for SARS-CoV-2 monitoring, it is key to understand the feasibility of using both alternative swabs and sample storage solutions. Here we provide evidence that the variation observed in a given SARSs-CoV-2 experiment is primarily driven by the time and method of sample collection rather than by the swab type, storage solution, and subsequent extraction and RT-qPCR. When using alcohol-based storage solutions, we demonstrate that the nucleic acid or viral particles tend to become enriched on the swab head rather than the eluent and thus we recommend extracting directly from the swab head itself. We demonstrate that RNA can be successfully extracted from consumer-grade swabs stored in alcohol without compromising RNA integrity or yield. Of note, wooden-shafted swabs performed poorly only when extracting from the eluent, suggesting that RNA adsorption onto the shaft, rather than RT-qPCR inhibitors, may be the source of interference with current eluent-based testing methods for this swab type. As cotton-tipped swabs and alcohol-based storage solutions are compatible with standard microbiome- and metabolome analyses not feasible with VTM, these alternatives could enable more widespread assessment of the microbial context of SARS-CoV-2 RNA in human and environmental samples, including associated microbiome features.

We also provide preliminary evidence that nasal samples collected using more widely available, consumer-grade, cotton-tipped swabs can be used to detect SARS-CoV-2 in the clinical setting. As cotton-tipped TMI swabs had only a marginally reduced performance compared to CDC-compliant PE swabs for nasal samples compared to NP results, these swabs provide the potential as an attractive alternative for methods such as metabolomics that are complicated by the background from incompatible with polyester-tipped swabs, as well as suggesting the expanding the pool of available medical-grade collection consumables could be expanded. Notably, this variation is less than that observed when comparing different methods for assessing the presence of SARS-CoV-2. Larger-scale testing will be needed to expand and confirm these findings, but our data suggests that these two swab types, in either 95% EtOH or isopropanol, would provide a valuable starting point.

When considering environmental sampling, our data suggest that TMI and CGp swabs may outperform or at least are similar to, CDC-compliant PE swabs for collecting samples to detect SARS-CoV-2 from floor samples. We provide molecular evidence demonstrating the feasibility of detecting SARs-CoV-2 from floor samples with a limit of detection (*ca*. 362.5 copies per extraction for TMI) and (750-1450 copies per extraction for CGp and PE) similar to that of other published studies (500 copies per extraction) [24]. Additional testing using pre-wetted swab heads, as performed in other built-environment studies [25–29], is warranted to determine if this would improve the ability of all swab types to detect SARS-CoV-2 in the hospital room environment. The detection of SARS-CoV-2 on *ca*. 50% of COVID-19(+) participants’ bedrails and *ca*. 75% of floors, as well as the detection of SARS-CoV-2 on the floor of one non-COVID patient, suggests increased cleaning measures may need to be taken. Indeed, the floor may be a potentially important reservoir for viral exposure, as shoe-covers are not currently recommended by the CDC. However, additional testing is needed to determine whether viable virus particles remain on these surfaces. Since detection largely does not differ across swab types, this suggests that differences seen in the quantitation of SARSs-CoV-2 in the clinic in a given floor or nasal sample is due to variation in the swabbing event itself rather than a molecular processing problem. Because of this, we recommend the need for standardization in medical devices used to collect both nasal and environmental samples specific to SARSs-CoV-2 to improve overall accuracy. Lastly, our efforts to quantify the total noise in a given sampling event and sample processing itself demonstrate how variation in the act of swabbing combined with sample processing may lead to variance and at times lower than expected specificity.

Secondary infections are an important and significant contributing factor to morbidity and mortality in COVID-19 patients [30,31]. With metagenomics assays becoming more common for infectious disease diagnostics in the clinic [32–34], developing molecular methods which enable simultaneous viral detection and metagenomic analysis is critical for understanding disease progression in at-risk populations. Since the storage method is a critical step in preserving microbiome integrity with 95% ethanol as a stable solution [35], our results further demonstrate and open the door for multi-omics processing and analysis of SARSs-CoV-2 samples.

In summary, our results suggest detection of SARS-CoV-2 RNA in the environment could be performed using less expensive, consumer-grade materials and alcohol-based storage solutions. With the materials examined in this study, it is further conceivable that patients could collect samples from themselves, their environments at home, or their place of work, dramatically expanding the ability to deploy widespread methods for monitoring and predicting outbreak events. Additional confirmatory studies using consumer-grade swabs would greatly support COVID-19 screening worldwide, particularly in resource-limited communities.

## Data Availability

16S rRNA sequencing data is available on Qiita (qiita.ucsd.edu) at Study ID 13275. qPCR data will be made available upon reasonable request.

## Acknowledgements

We thank research participants who donated samples, the health care professionals who assisted in the collection of samples, and Alison Vrbanac, Louis-Felix Nothias-Scaglia, and Shi Huang for assistance in transportation and Dominic Nguyen for sampling kit preparation. This research benefited tremendously from lessons learned and techniques developed in the Sloan Microbiology of the Built Environment (MoBE program). We thank Stanley T. Motley for early discussion on nucleic acid stability, Sandrine Miller-Montgomery for guidance in RT-qPCR analyses. We thank Thomas F. Rogers and Nathan Beutler for providing the SARS-CoV-2 viral RNA used as a control in this study and David Pride for supplying samples from NP swabs in VTM for our comparisons.

This work was supported by the UC San Diego Center for Microbiome Innovation (CMI). JJM is supported by the National Science Foundation Center for Aerosol Impacts on Chemistry of the Environment #1801971. CM is supported by NIDCR NRSA F31 Fellowship 1F31DE028478-01. PBF is partially funded through trainee support from Sanitarium Health and Wellbeing in partnership with the CMI. DM is partially funded by support from Danone Nutricia Research in partnership with the CMI. JPS is supported by NIH-SD-IRACDA (5K12GM068524-17) and USDA-NIFA (2019-67013-29137).

## Supplementary Figures and Legends

**Supplemental Table 1.**

| Supplemental Table. SARs-CoV-2 Limit of detection: total quantified copies of SARs-CoV-2 (N1 or N2) and human Rp across swab types. |  |  |  |  |  |  |  |  |  |  |  |  |
| --- | --- | --- | --- | --- | --- | --- | --- | --- | --- | --- | --- | --- |
|  | N1 |  |  |  |  |  |  |  |  |  |  |  |
|  | noswab |  |  | PE |  |  | CGp |  |  | TMI |  |  |
| 0 | 0 | 0 | 0 | 0 | 0 | 0 | 0 | 0 | 0 | 0 | 0 | 0 |
| 362.5 | 1683 | 749 | 782 | 866 | 0 | 203 | 0 | 600 | 1539 | 195 | 146 | 173 |
| 725 | 420 | 599 | 713 | 0 | 2213 | 752 | 1585 | 1175 | 0 | 1204 | 859 | 755 |
| 1450 | 1704 | 1722 | 1665 | 209 | 946 | 2411 | 1390 | 1618 | 2572 | 1021 | 3147 | 1071 |
| 2900 | 4305 | 3868 | 3157 | 1432 | 2801 | 4794 | 4053 | 2834 | 2153 | 2166 | 4646 | 4533 |
| 5800 | 7598 | 6897 | 8862 | 3819 | 8958 | 5086 | 5836 | 10027 | 7527 | 12332 | 10290 | 10227 |
| 11600 | 19177 | 14181 | 13316 | 11752 | 11062 | 11885 | 20925 | 10598 | 13416 | 17761 | 18618 | 15658 |
| 116000 | 138318 | 121286 | 151498 | 104301 | 139976 | 94119 | 163019 | 111423 | 138201 | 212959 | 233003 | 172233 |
|  | N2 |  |  |  |  |  |  |  |  |  |  |  |
|  | noswab |  |  | PE |  |  | CGp |  |  | TMI |  |  |
| 0 | 0 | 0 | 0 | 0 | 0 | 0 | 0 | 0 | 0 | 0 | 0 | 0 |
| 362.5 | 218 | 0 | 133 | 419 | 0 | 125 | 0 | 0 | 643 | 420 | 131 | 289 |
| 725 | 719 | 474 | 345 | 308 | 458 | 127 | 217 | 222 | 0 | 356 | 484 | 489 |
| 1450 | 263 | 1356 | 371 | 170 | 345 | 1099 | 1550 | 633 | 889 | 1023 | 1626 | 61 |
| 2900 | 1536 | 2525 | 781 | 622 | 1004 | 1228 | 1694 | 1289 | 937 | 1587 | 1146 | 1371 |
| 5800 | 2350 | 2764 | 3261 | 2718 | 3527 | 1733 | 2924 | 6110 | 2246 | 4861 | 5149 | 3254 |
| 11600 | 5230 | 5572 | 6646 | 5569 | 4899 | 5883 | 6011 | 3656 | 4922 | 8808 | 8611 | 7388 |
| 116000 | 49810 | 53165 | 57606 | 43923 | 66597 | 43910 | 46084 | 41405 | 50328 | 93101 | 102074 | 61855 |
|  | Rp |  |  |  |  |  |  |  |  |  |  |  |
|  | noswab |  |  | PE |  |  | CGp |  |  | TMI |  |  |
| 0 | 0 | 0 | 0 | 521 | 462 | 391 | 347 | 86 | 66 | 0 | 79 | 0 |
| 362.5 | 0 | 0 | 0 | 1861 | 737 | 283 | 164 | 263 | 185 | 130 | 83 | 0 |
| 725 | 0 | 0 | 0 | 723 | 3368 | 921 | 67 | 81 | 0 | 207 | 0 | 0 |
| 1450 | 0 | 0 | 0 | 283 | 426 | 499 | 0 | 0 | 377 | 135 | 198 | 147 |
| 2900 | 0 | 0 | 0 | 1254 | 479 | 222 | 194 | 97 | 422 | 0 | 156 | 0 |
| 5800 | 0 | 0 | 0 | 252 | 873 | 491 | 0 | 220 | 0 | 97 | 0 | 0 |
| 11600 | 0 | 0 | 0 | 149 | 526 | 1112 | 203 | 426 | 220 | 52 | 77 | 143 |
| 116000 | 0 | 0 | 0 | 330 | 798 | 1047 | 3980 | 0 | 256 | 117 | 97 | 0 |

**Supplemental Figure 1.**
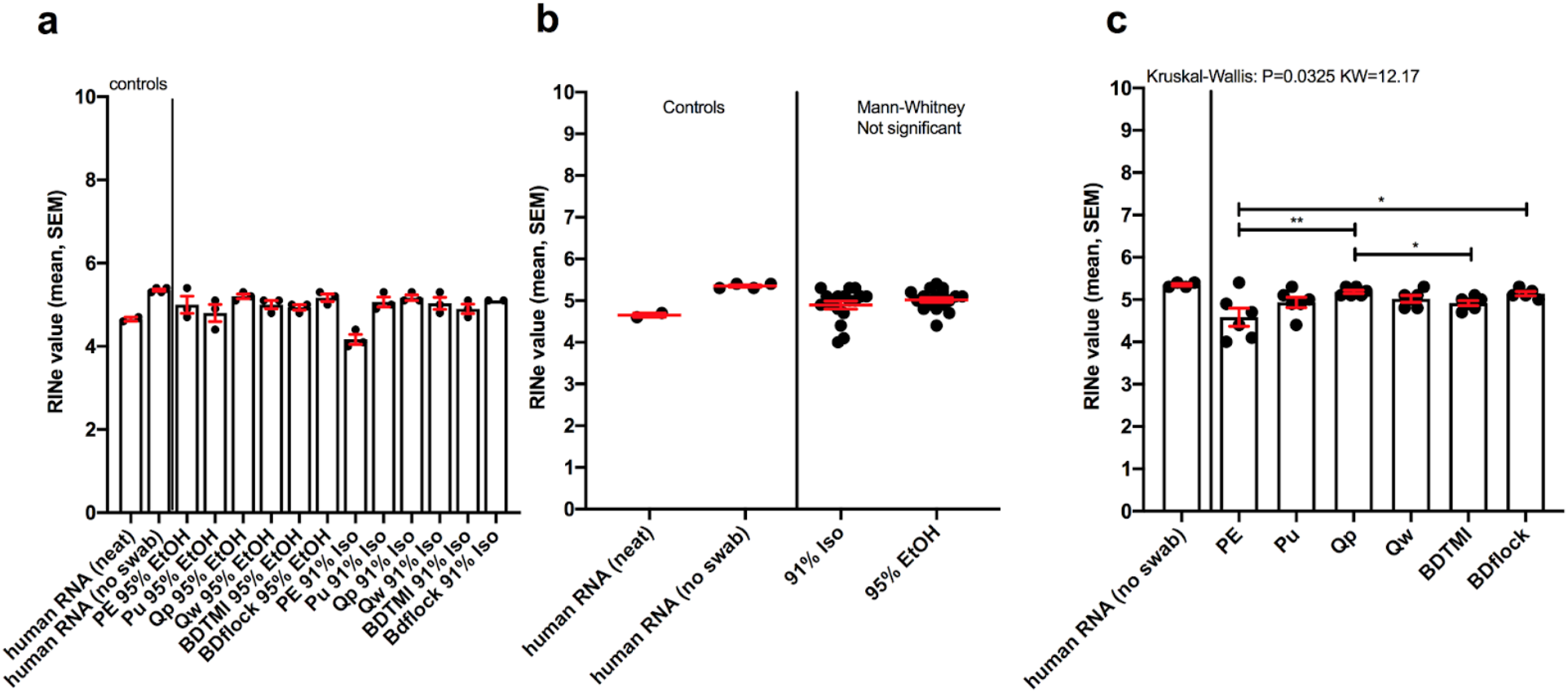
Impacts of storage solution or swab type on RNA quality as measured by RNA Tapestation High Sensitivity kit. a) All direct-swab extracted RNA grouped by storage buffer and swab type. b) Samples grouped by storage buffer, no significant difference in RNA integrity number (RIN) values between storage buffers (95% EtOH vs. 91% isopropanol) (Mann-Whitney). c) Swab extracts grouped by swab type only and compared to determine if swab type has an impact on RNA quality (Kruskal-Wallis test).

**Supplemental Figure 2.**
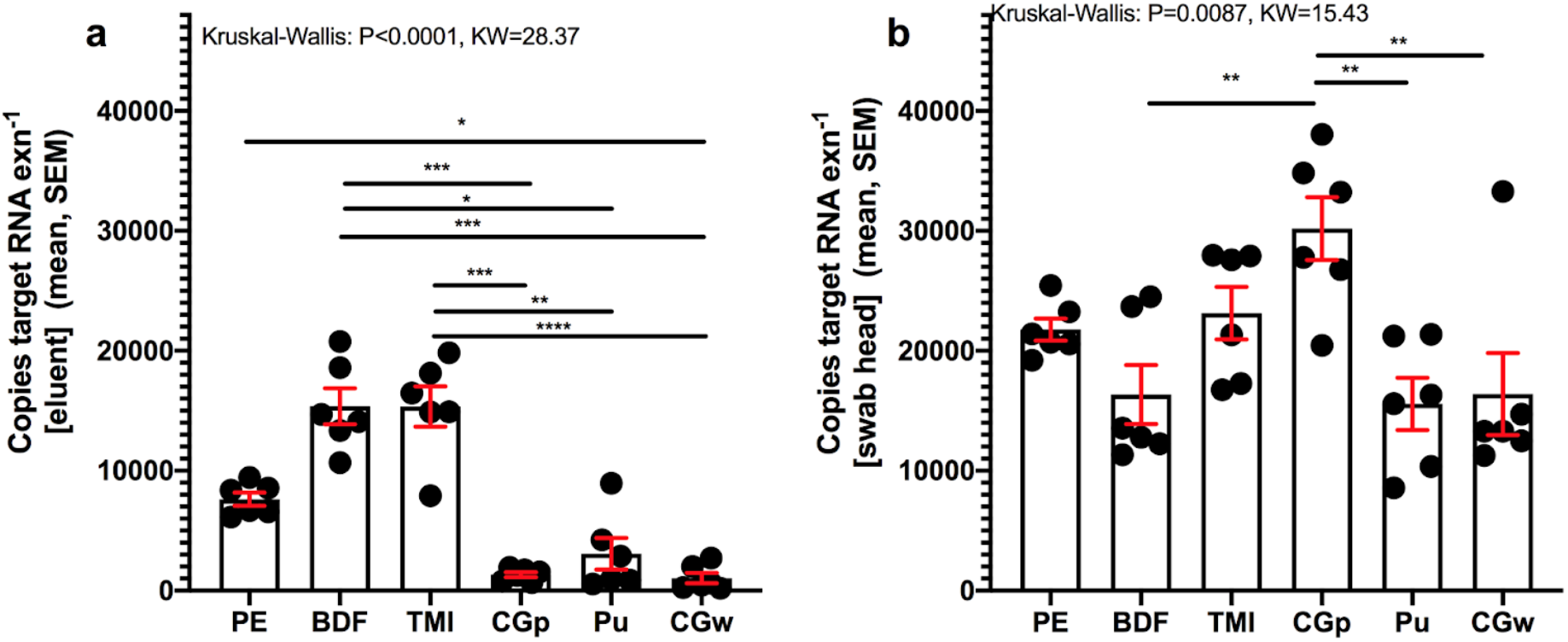
Impacts of sample-type (eluent vs. swab head) on RNA recovery by swab used. Comparison human RNA recovery across six swab types (PE=polyester ‘commercial’, BDF=BD foam ‘commercial’, TMI=BD TMI ‘commercial’, CGp=plastic ‘consumer-grade’, Pu=Puritan ‘commercial’, CGw=wood ‘consumer-grade’), **a)** extracted from 200 μL eluent or **b)** swab head (Group comparison using Kruskal-Wallis)

**Supplemental Figure 3.**
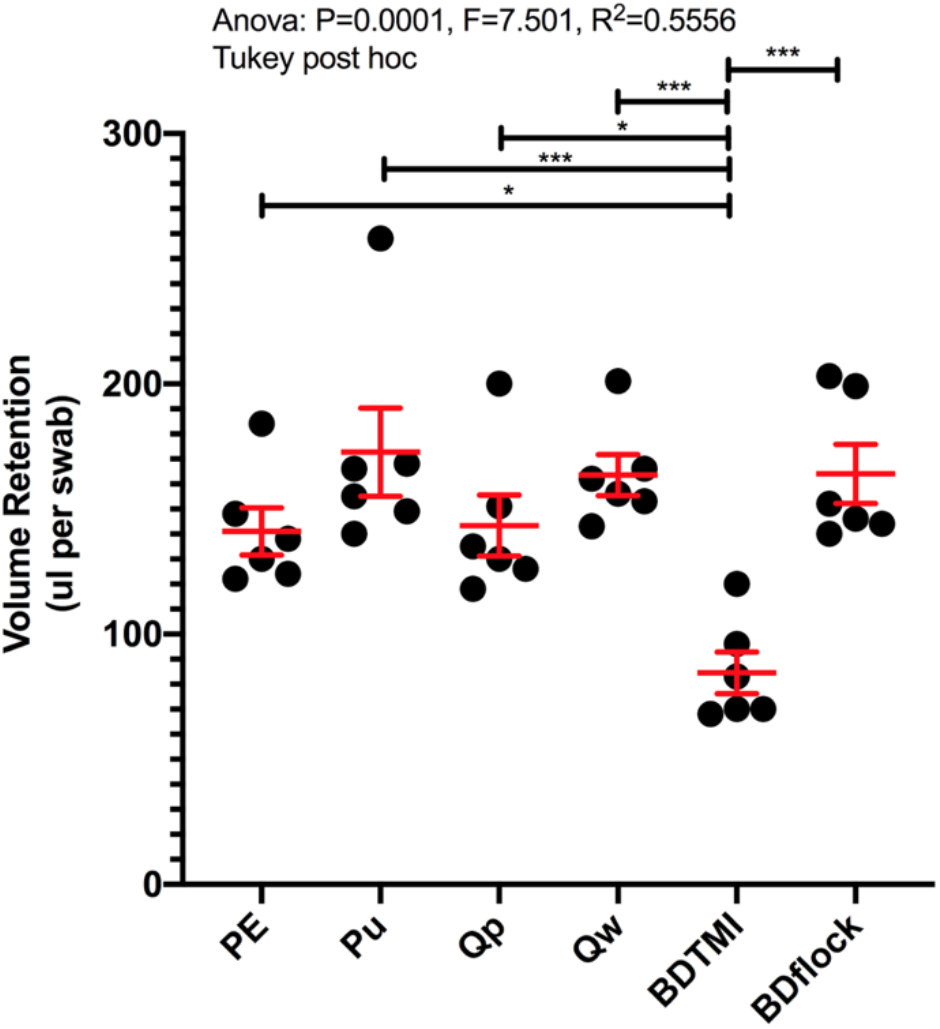
Adsorption volumes per swab type. Samples tested for normality using Shapiro-Wilke test (not significant, P=0.12, W=0.8357).

**Supplemental Figure 4.**
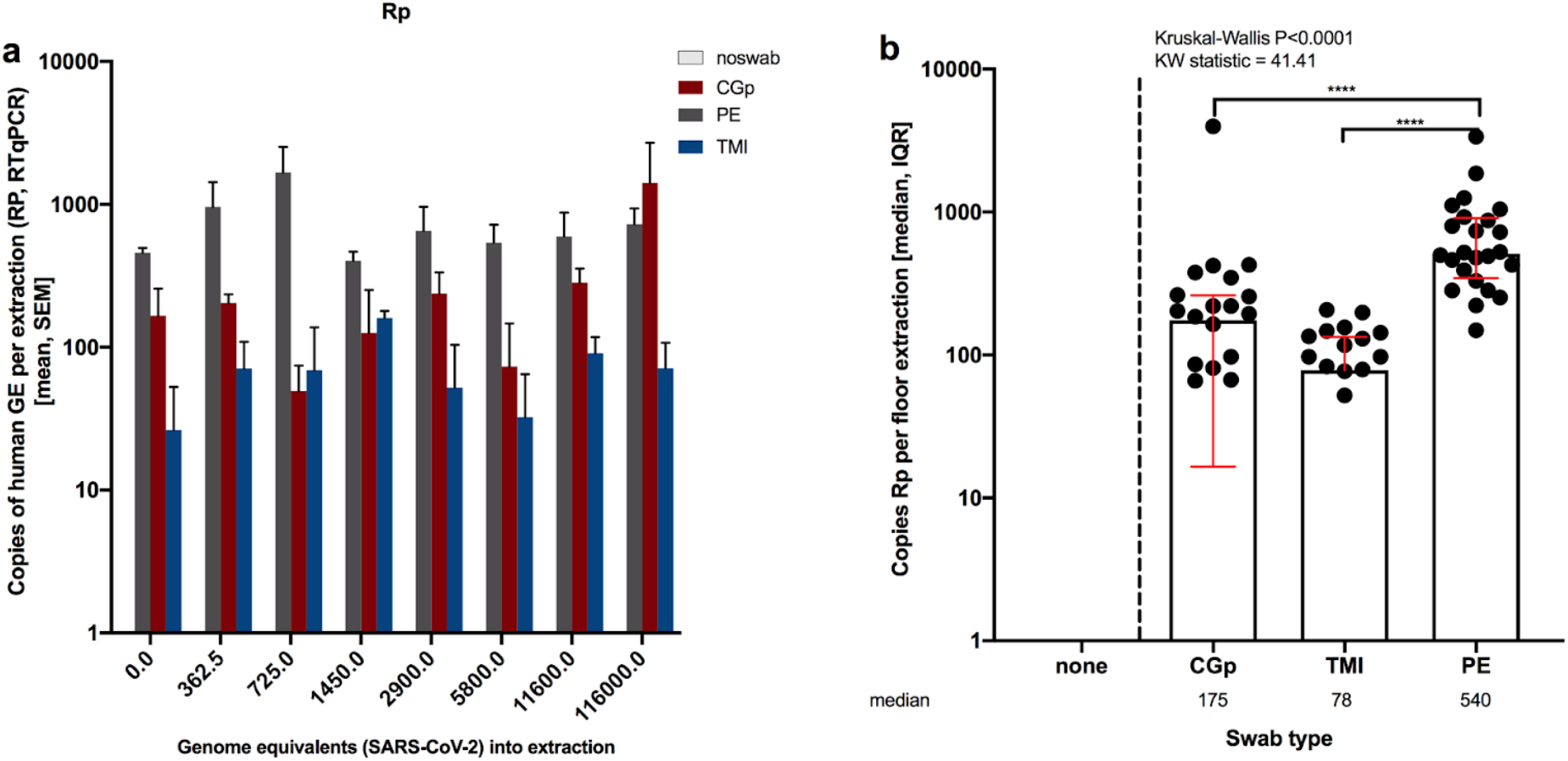
Total variation of human Rp gene detected from floor samples in limit of detection experiment. a) Variation of Rp across swab types and subsequent dilutions. b) total yield of human Rp across aggregated swab types.

**Supplemental Figure 5.**
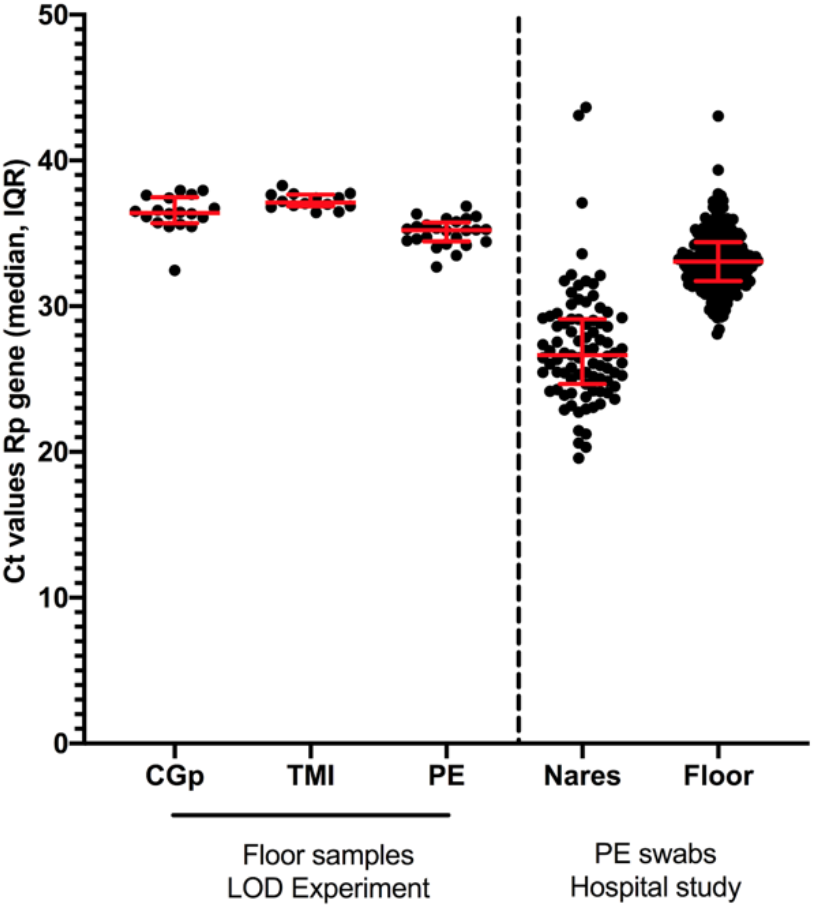
Variation of human Rp gene amplification across swab types. (left) Surface swabs were collected from the floor of a laboratory to compare efficiency across swab types; consumer-grade cotton (CGp), The Microsetta Initiative cotton swabs (TMI), and polyester (PE). (right) PE surface swab samples were collected with the same protocol in the hospital. The variation across hospital samples is greater than the variation among swab types.

**Supplemental Figure 6.**
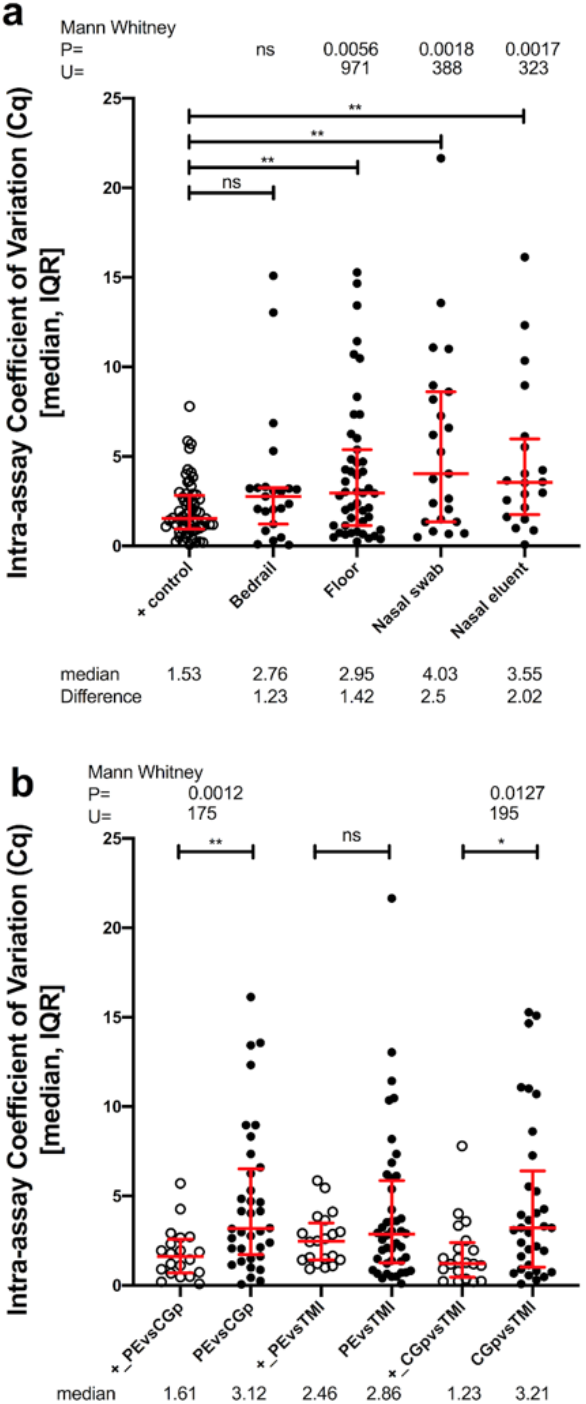
Intra-assay variability compared across positive controls, sample _types, and swab types. **a)** Coefficient of variance calculated across pairwise swabs taken within each room/patient replicate and then distributions compared for bedrail, floor, nasal swab, and nasal eluent samples along with positive controls. b) The CV was also grouped and compared by swab type.

**Supplemental Figure 7.**
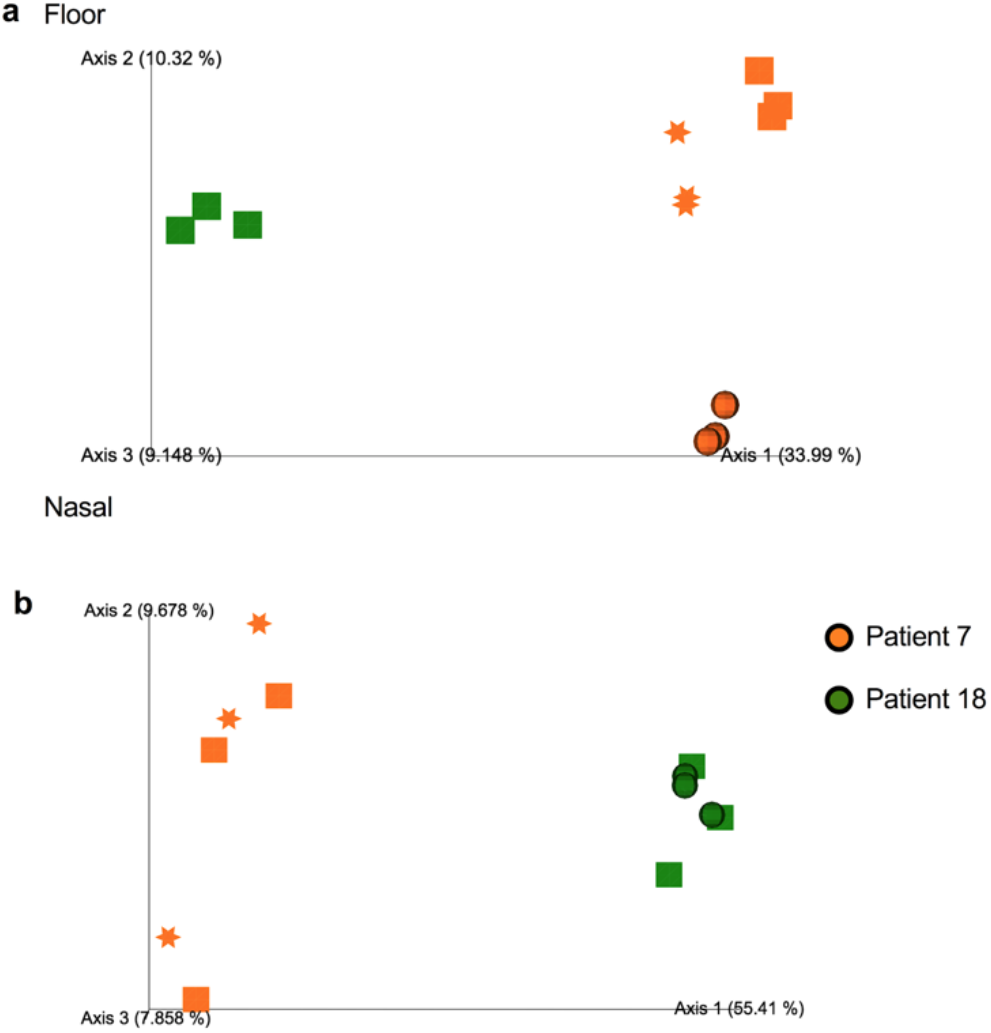
Comparison of 16S rRNA microbial composition of paired patient and sample type samples collected using different swabs (in triplicate) using Unweighted UniFrac PCoA plot. a) floor samples, b) nasal samples. Swab types: square = PE, circle = CGp, star = TMI.

## References

1. Mediterranean 828 Deaths Eastern, Cases 98, Europe 664 Deaths, Cases 2. 827 789, Pacific 224 Deaths Western, Cases 31 749, et al. Situation in numbers (by WHO Region). Available from: https://www.who.int/docs/default-source/coronaviruse/situation-reports/20200708-covid-19-sitrep-170.pdf?sfvrsn=bca86036_2

2. Gilbert JA, Stephens B. Microbiology of the built environment. Nat Rev Microbiol. 2018;16:661–70.

3. CDC Background F. Environmental Sampling [Internet]. 2019 [cited 2020 Jul 9]. Available from: https://www.cdc.gov/infectioncontrol/guidelines/environmental/background/sampling.html

4. Centers for Disease Control and Prevention. Centers for Disease Control and Prevention SOP#: DSR-052-02-PREPARATION OF VIRAL TRANSPORT MEDIUM [Internet]. Centers for Disease Control and Prevention-PREPARATION OF VIRAL TRANSPORT MEDIUM. 2020 [cited 2020 Apr 24]. Available from: https://www.cdc.gov/coronavirus/2019-ncov/downloads/Viral-Transport-Medium.pdf

5. Kalantar-Zadeh K, Ward SA, Kalantar-Zadeh K, El-Omar EM. Considering the Effects of Microbiome and Diet on SARS-CoV-2 Infection: Nanotechnology Roles. ACS Nano [Internet]. 2020; Available from: http://dx.doi.org/10.1021/acsnano.0c03402

6. Bergner LM, Orton RJ, da Silva Filipe A, Shaw AE, Becker DJ, Tello C, et al. Using noninvasive metagenomics to characterize viral communities from wildlife. Mol Ecol Resour. 2019; 19:128–43.

7. Oropharyngeal 1., Swabs N. Collection of Upper Respiratory Tract Specimens. Available from: https://www.cdc.gov/urdo/downloads/SpecCollectionGuidelines.pdf

8. CDC/DDID/NCIRD/ Division of Viral Diseases. CDC 2019-Novel Coronavirus (2019-nCoV) Real-Time RT-PCR Diagnostic Panel. 2020; Available from: https://www.fda.gov/media/134922/download

9. Marx V. Coronavirus jolts labs to warp speed. Nat Methods. 2020;17:465–8.

10. Minich JJ, Zhu Q, Janssen S, Hendrickson R, Amir A, Vetter R, et al. KatharoSeq Enables High-Throughput Microbiome Analysis from Low-Biomass Samples. mSystems [Internet]. 2018;3. Available from: http://dx.doi.org/10.1128/mSystems.00218-17

11. CDC. Coronavirus Disease 2019 (COVID-19) [Internet]. Centers for Disease Control and Prevention. 2020 [cited 2020 Apr 21]. Available from: https://www.cdc.gov/coronavirus/2019-ncov/lab/rt-pcr-panel-primer-probes.html

12. Minich JJ, Humphrey G, Benitez RAS, Sanders J, Swafford A, Allen EE, et al. High-Throughput Miniaturized 16S rRNA Amplicon Library Preparation Reduces Costs while Preserving Microbiome Integrity. mSystems [Internet]. 2018;3. Available from: http://dx.doi.org/10.1128/mSystems.00166-18

13. Apprill A, McNally S, Parsons R, Weber L. Minor revision to V4 region SSU rRNA 806R gene primer greatly increases detection of SAR11 bacterioplankton [Internet]. Aquatic Microbial Ecology. 2015. p. 129–37. Available from: http://dx.doi.org/10.3354/ame01753

14. Caporaso JG, Lauber CL, Walters WA, Berg-Lyons D, Lozupone CA, Turnbaugh PJ, et al. Global patterns of 16S rRNA diversity at a depth of millions of sequences per sample [Internet]. Proceedings of the National Academy of Sciences. 2011. p. 4516–22. Available from: http://dx.doi.org/10.1073/pnas.1000080107

15. Walters W, Hyde ER, Berg-Lyons D, Ackermann G, Humphrey G, Parada A, et al. Improved Bacterial 16S rRNA Gene (V4 and V4-5) and Fungal Internal Transcribed Spacer Marker Gene Primers for Microbial Community Surveys. mSystems [Internet]. 2016; 1. Available from: http://dx.doi.org/10.1128/mSystems.00009-15

16. Parada AE, Needham DM, Fuhrman JA. Every base matters: assessing small subunit rRNA primers for marine microbiomes with mock communities, time series and global field samples. Environ Microbiol. 2016;18:1403–14.

17. Gonzalez A, Navas-Molina JA, Kosciolek T, McDonald D, Vázquez-Baeza Y, Ackermann G, et al. Qiita: rapid, web-enabled microbiome meta-analysis. Nat Methods. 2018;15:796–8.

18. Estaki M, Jiang L, Bokulich NA, McDonald D, González A, Kosciolek T, et al. QIIME 2 Enables Comprehensive End-to-End Analysis of Diverse Microbiome Data and Comparative Studies with Publicly Available Data. Curr Protoc Bioinformatics. 2020;70:e100.

19. Bolyen E, Rideout JR, Dillon MR, Bokulich NA, Abnet CC, Al-Ghalith GA, et al. Reproducible, interactive, scalable and extensible microbiome data science using QIIME 2. Nat Biotechnol. 2019;37:852–7.

20. Amir A, McDonald D, Navas-Molina JA, Kopylova E, Morton JT, Zech Xu Z, et al. Deblur Rapidly Resolves Single-Nucleotide Community Sequence Patterns. mSystems [Internet]. 2017;2. Available from: http://dx.doi.org/10.1128/mSystems.00191-16

21. Vazquez-Baeza Y, Pirrung M, Gonzalez A, Knight R. EMPeror: a tool for visualizing high-throughput microbial community data. Gigascience. 2013;2:16.

22. Schroeder A, Mueller O, Stocker S, Salowsky R, Leiber M, Gassmann M, et al. The RIN: an RNA integrity number for assigning integrity values to RNA measurements. BMC Mol Biol. 2006;7:3.

23. Zou L, Ruan F, Huang M, Liang L, Huang H, Hong Z, et al. SARS-CoV-2 Viral Load in Upper Respiratory Specimens of Infected Patients. N Engl J Med. 2020;382:1177–9.

24. Vogels CBF, Brito AF, Wyllie AL, Fauver JR, Ott IM, Kalinich CC, et al. Analytical sensitivity and efficiency comparisons of SARS-CoV-2 RT-qPCR primer-probe sets. Nat Microbiol [Internet]. 2020; Available from: http://dx.doi.org/10.1038/s41564-020-0761-6

25. Lax S, Cardona C, Zhao D, Winton VJ, Goodney G, Gao P, et al. Microbial and metabolic succession on common building materials under high humidity conditions. Nat Commun. 2019; 10:1767.

26. Richardson M, Gottel N, Gilbert JA, Lax S. Microbial Similarity between Students in a Common Dormitory Environment Reveals the Forensic Potential of Individual Microbial Signatures. MBio [Internet]. 2019; 10. Available from: http://dx.doi.org/10.1128/mBio.01054-19

27. Lax S, Sangwan N, Smith D, Larsen P, Handley KM, Richardson M, et al. Bacterial colonization and succession in a newly opened hospital. Sci Transl Med. 2017;9.

28. Wood M, Gibbons SM, Lax S, Eshoo-Anton TW, Owens SM, Kennedy S, et al. Athletic equipment microbiota are shaped by interactions with human skin. Microbiome. 2015;3:25.

29. Lax S, Smith DP, Hampton-Marcell J, Owens SM, Handley KM, Scott NM, et al. Longitudinal analysis of microbial interaction between humans and the indoor environment. Science. 2014;345:1048–52.

30. Alhazzani W, Møller MH, Arabi YM, Loeb M, Gong MN, Fan E, et al. Surviving Sepsis Campaign: Guidelines on the Management of Critically Ill Adults with Coronavirus Disease 2019 (COVID-19). Crit Care Med. 2020;48:e440–69.

31. Ruan Q, Yang K, Wang W, Jiang L, Song J. Clinical predictors of mortality due to COVID-19 based on an analysis of data of 150 patients from Wuhan, China. Intensive Care Med. 2020;46:846–8.

32. Deng X, Achari A, Federman S, Yu G, Somasekar S, Bártolo I, et al. Metagenomic sequencing with spiked primer enrichment for viral diagnostics and genomic surveillance. Nat Microbiol. 2020;5:443–54.

33. Wilson MR, Sample HA, Zorn KC, Arevalo S, Yu G, Neuhaus J, et al. Clinical Metagenomic Sequencing for Diagnosis of Meningitis and Encephalitis. N Engl J Med. 2019;380:2327–40.

34. Chiu CY, Miller SA. Clinical metagenomics. Nat Rev Genet. 2019;20:341–55.

35. Song SJ, Amir A, Metcalf JL, Amato KR, Xu ZZ, Humphrey G, et al. Preservation Methods Differ in Fecal Microbiome Stability, Affecting Suitability for Field Studies. mSystems [Internet]. 2016; 1. Available from: http://dx.doi.org/10.1128/mSystems.00021-16

